# Regorafenib in Bone Sarcoma: A Systematic Review and Meta-Analysis of Its Effectiveness and Safety in Cancer Treatment

**DOI:** 10.1101/2025.08.04.25332569

**Authors:** Mohsen Rahmanian, Sarah Khoropanah, Abulfazl Vatankhah, Sepehr Hosseinzadeh Moghaddam, Elaheh Abdi Bastami, Soheila Roashanzamir, Amir Rahmanian Sharifabad, Reza Ganji

## Abstract

**Objectives:** Bone sarcomas are aggressive malignancies with limited treatment options in advanced stages. Regorafenib, an oral multi-kinase inhibitor, has showed potential efficacy in early-phase trials. However, its overall effectiveness and safety profile in bone sarcoma remain unclear. This systematic review and meta-analysis aim to assess the clinical impact of regorafenib on survival outcomes and adverse events in patients with bone sarcomas.

**Methods:** A systematic search was conducted across PubMed, Web of Science, and Scopus for randomized controlled trials (RCTs) published between September 27, 2012, and April 2024. Eligible studies included full-text RCTs comparing regorafenib to placebo in the treatment of bone sarcomas. Meta-analyses, reviews, case reports, conference abstracts, ongoing trials, and animal studies were excluded from the analysis. The primary endpoints were progression-free survival (PFS), overall survival (OS), and adverse events (AEs). Data synthesis was performed using a random-effects model, and heterogeneity was assessed via the I² statistic. The Cochrane Risk of Bias tool (RoB 2) was applied to evaluate study quality. Statistical analysis was conducted using R version 4.3.1.

**Results:** A total of five RCTs met the inclusion criteria. Regorafenib significantly improved PFS compared to placebo (MD = 9.69 weeks; 95% CI: 4.54, 14.84; I² = 0%), while no statistically significant improvement was observed in OS (MD = 0.85 weeks; 95% CI:-36.33, 38.02; I² = 0%). Predominantly observed treatment-emergent effects linked to regorafenib included hand-foot skin reaction, hypertension, asthenia or fatigue, and diarrhea.

**Conclusion:** Regorafenib significantly improves progression-free survival in bone sarcoma but does not show a clear benefit in overall survival. While adverse events are common, they are generally manageable. Further research is needed to optimize dosing, patient selection, and to determine the long-term survival benefits.

## 1. Introduction

Bone sarcomas comprise 0.2% of U.S. cancer cases annually, with 3,300 new cases in 2016, rising 0.4% yearly. (^1^) It can be classified into several types, including chondrosarcoma, Ewing sarcoma, and osteosarcoma, as well as rarer conditions such as chordoma and adamantinoma. (^2^) Overall treatment strategy should depend on a multitude of factors. (^3^) A priority hierarchy in bone sarcoma care includes life, limb, function, length equality, and cosmesis. Surgical resection ensures local control, while chemotherapy provides systemic control for eligible histologies. Limb salvage surgery is increasingly favored over amputation to enhance function without compromising survival. (^4,5^)

Chemotherapy for adult bone sarcoma is challenging, with varying responses. Conventional chondrosarcoma shows minimal response, while mesenchymal and dedifferentiated subtypes may respond better to specific treatments. (^6,7^) The effectiveness of chemotherapy in managing metastatic bone sarcoma is generally limited. (^8^) Additionally, older age is often correlated with poorer treatment outcomes. (^6^) Moreover, the adverse effects of chemotherapy, including nausea, vomiting, alopecia, fatigue, and heightened infection risk, pose challenges and can affect the quality of life. (^9^)

Numerous treatments have been introduced in oncology because of the limitations of conventional therapies. Tyrosine kinase inhibitors represent a new class of drugs for treating cancers with poor prognosis, and various clinical trials are investigating their efficacy in bone sarcoma. (^10,11^) Regorafenib is one of the emerging drugs in the tyrosine kinase inhibitors class which suppresses the function of angiogenic, stromal, and oncogenic receptor tyrosine kinases. (^12,14–16^) This paper aims to evaluate the use of regorafenib in the treatment of bone sarcoma and to determine its potential as a viable treatment option for this challenging disease.

## 2. Materials and Methods

### 2.1. Search Strategy

An extensive literature exploration identified relevant published articles on randomized clinical trials (RCTs) from September 27, 2012, to April 2024. The search was conducted in PubMed, Web of Science, and Scopus. The main keywords used for the search were ((Bone sarcoma) OR (Chondrosarcoma) OR (chondroblastoma) OR (chordoma) OR (Ewing sarcoma) OR (osteosarcoma) OR (bone fibrosarcoma) OR (bone leiomyosarcoma) OR (Chordoma) OR (Adamantinoma) OR (Hemangioendothelioma) OR (Hemangiopericytoma) OR (Low-Grade Fibrosarcoma) OR (Malignant Fibrous Histiocytoma)) AND (Stivarga OR regorafenib).

This review protocol has been formally documented in the International Prospective Register of Systematic Reviews (PROSPERO) under ID CRD42024526345. Additionally, the present investigation adhered to the 2020 guidelines of the Preferred Reporting Items for Systematic Reviews and Meta-Analyses (PRISMA). An initial selection was performed using citation titles and abstracts, followed by a detailed evaluation of full-text articles to determine their suitability for inclusion in the review.

### 2.2. Eligibility Criteria

This analysis was limited to Randomized Controlled Trials (RCTs) assessing regorafenib’s efficacy in the treatment of bone sarcoma. The location and language of the studies were not a factor in the selection process. However, only studies with full-text results published were considered for inclusion. Furthermore, all included studies must have examined the efficacy and safety of regorafenib with a placebo in treating bone sarcoma.

During the screening process, one author (M.R.) examined the titles and abstracts of records identified through the literature search. In the initial search and selection process, two independent authors (M.R. and S.K.) reviewed the full texts of the articles. In the event of disagreements, an open discussion was held among the authors to reach a consensus. If the difference of opinion persisted, a third author (A.T.), acting as a referee, adjudicated the debate to maintain the integrity of the review process.

The inclusion date criteria were from September 27, 2012, marking the date when regorafenib was approved by the FDA for patients, until the completion of the literature search. This period was chosen to provide a comprehensive overview of the developments and findings in the field since the introduction of regorafenib to clinical practice.

Studies were excluded from the review if they were meta-analyses, literature reviews, case reports, abstracts, guidelines, ongoing trials, trial protocols, Letters to the Editor, or animal studies, or if they lacked the data necessary to evaluate the primary and secondary outcomes or provided only a partial PDF text.

### 2.3. Data Extraction

Information and results were gathered from the selected studies using standardized digital formats created beforehand by two independent authors (M.R. and S.K.). In case of disagreements, the authors discussed reaching a consensus. If unresolved, a third author (S.H.) acted as a referee. The data points obtained in this manner comprised author names, years, countries, the studied populations, type of clinical trial, participants’ initial characteristics, exact histopathological sarcoma type, recruitment period, Progression-Free Rate (PFR), Response Rate (RR), and Overall Survival (OS). Participant characteristics, such as the male-to-female ratio, age, and other relevant information, were also compiled from the available research at the start of the studies. Treatment-related adverse effects (AEs) provided necessary measures to complement outcome data. Where specific data points for the outcomes of interest (PFS, OS, AEs) were not reported thoroughly, we contacted the corresponding authors for clarification. However, if no response was received, the study was excluded from the analysis of that specific outcome.

### 2.4. Synthesis methods and Statistical analysis

This pooled analysis assessed PFS, OS, and AEs, including studies with sufficient data. Inverse variance weighting was applied, with mean difference (MD) in weeks for PFS/OS and odds ratio (OR) for AEs. Outcomes were standardized across studies. Heterogeneity (I² statistic) was categorized as high (>75%), moderate (50–75%), or low (25–50%), guiding the systematic analysis of treatment effects. (^17^) We used the random effects model because of possible wide heterogeneity in selected studies. Subgroup analyses were performed based on different histological subtypes of bone sarcoma. Additionally, tests for subgroup differences were conducted. Statistical significance was determined by a P value threshold set below 0.05. In this review, statistical computations were executed with R version 4.3.1.

## 3. Results

### 3.1. Study Selection

Through database searching, 400 potential literature were initially investigated. 101 publications were excluded due to duplication. 286 studies were further removed after the title/abstract screening. After a thorough examination of the full text, five articles were excluded because of the limited availability of the full text. In contrast, three articles were excluded because they lacked a control group. Finally, five articles were considered eligible for the final qualitative analysis. The flow diagram, which details the enrollment of the included literature, is shown in **[Table 1]**.

**Table 1.**
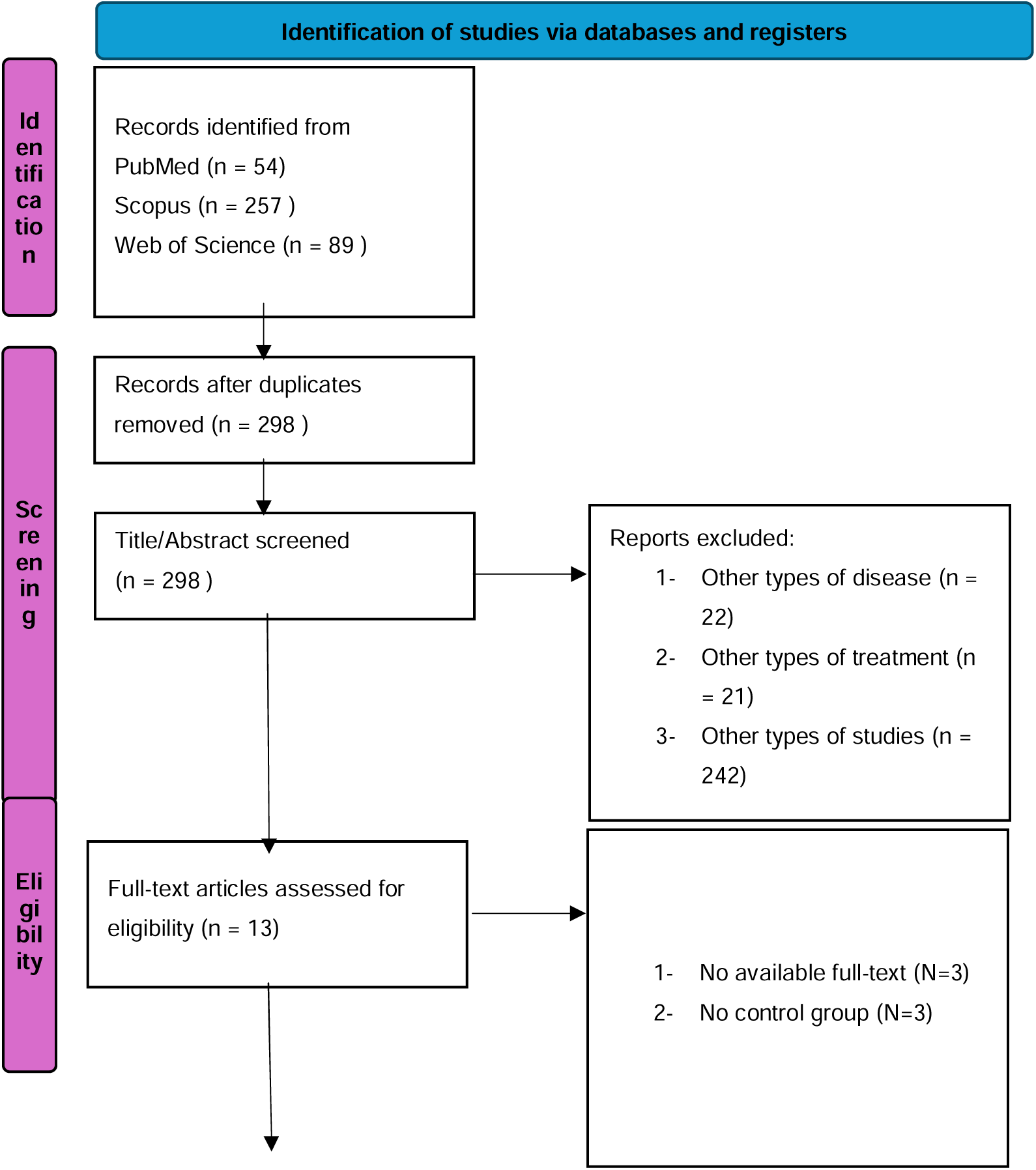
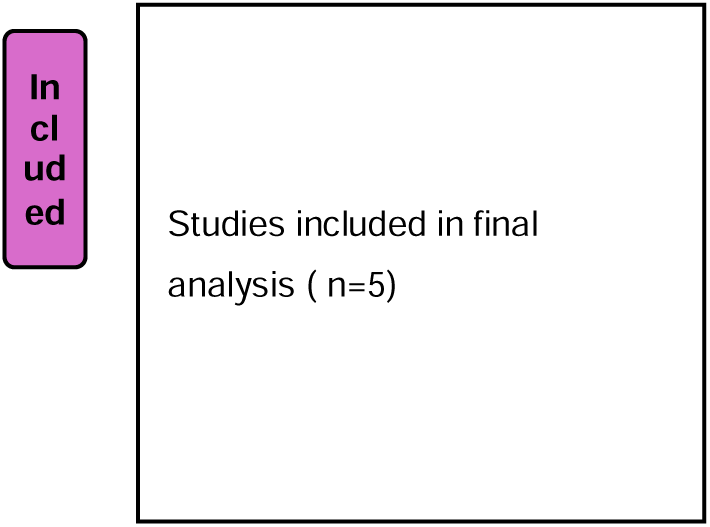
PRISMA Flow Diagram for Study Selection.

### 3.2 Study Characteristics

Five papers were incorporated into the evaluation. These studies were conducted as Phase II trials between 2019 and 2024. Main characteristics of trials is provided **[Table 2].**

**Table 2.** Characteristics of the included studies.

| Study | Year | Study phase | Design | Condition | Country | Recruitment period | Sample size | M/F | Age | Outcomes |
| --- | --- | --- | --- | --- | --- | --- | --- | --- | --- | --- |
| Davis *et al.* (18) | 2019 | Phase 2 | Randomized, double-blind, placebo-controlled | Osteosarcoma | USA | September 2014 - May 2018 | 42 patients - 22 in the regorafenib arm, 20 in the placebo arm | 20/22 | Median 37 years | Median PFS, OS, time to tumor progression, PFS at eight and 16 weeks, OS, Overall response rate, |
| Duffaud *et al.* (19) | 2019<sup>1</sup> | Phase 2 | Non-comparative, randomized, double-blind, placebo-controlled | Osteosarcoma | France | October 2014 - April 2017 | 43 patients - 29 in the regorafenib group, 14 in the placebo group | 24/14 | Median 33 years | Median PSF, OS, PFR at 8, 12, and 24 weeks, Adverse effects. |
| Duffaud *et al.* (20) | 2021 | Phase 2 | Non-comparative, randomized, double-blind, placebo-controlled | Chondrosarcoma | France | September 2014-February 2019 | 41 patients - 24 in the regorafenib group, 16 in the placebo group<sup>2</sup> | 25/15 | - | Median PSF, OS, PFR at 12 and 24 weeks, OS at six and 12 months, Adverse effects |

<sup>1</sup> This article was published online on November 23, 2018, and was subsequently included in the January 2019 issue of the journal
<sup>2</sup> One patient was excluded for efficacy analysis due to major protocol violation (modified intention-to-treat analysis); 25 patients was included in safety analysis.

|  |  |  |  |  |  |  |  |  |  |  |
| --- | --- | --- | --- | --- | --- | --- | --- | --- | --- | --- |
| Le Cesne *et al.*(21) | 2023 | Phase 2 | Randomized, double-blind, placebo-controlled | Chordoma | France | March 2016 - February 2020 | 27 patients total - 18 in the regorafenib arm, eight in the placebo arm | 16/7 | Median 66 years | PFR at six months, Median PFS, OS, response rate, objective response rate, duration of overall response, Adverse effects |
| Duffaud *et al.*(22) | 2023 | Phase 2 | Non-comparative, randomized, double-blind, placebo-controlled | Ewing sarcoma | France | September 24, 2014 to November 4, 2019 | 36 patients total- 23 in the regorafenib arm, 13 in the placebo arm | 28/8 | 32 in the regorafenib arm, 28 in the placebo arm | PFR at eight weeks, PFS Objective response rate (ORR), OS. Duration of response (DoR) Adverse effects |

### 3.3. Study Quality

The included studies in this study underwent an evaluation of methodological integrity through the Cochrane risk-of-bias instrument for randomized trials (RoB 2). Two independent authors (M.R. and S.H.) revised the studies, and in cases of disagreement, the authors discussed to reach a consensus. If differences persisted, a third author (A.T.) acted as a referee to resolve the issue **[Table 3]**.

**Table 3.** Risk of bias assessment for included studies.

| Study | D1: Randomization | D2: Intervention Deviation | D3: Missing Data | D4: Outcome Measurement | D5: Reported Result | Overall Risk |
| --- | --- | --- | --- | --- | --- | --- |
| Davis et al. 2019 | Some concerns | Low | Low | Low | Low | Some concerns |
| Duffaud et al. 2019 | Low | Low | Low | Low | Low | Low |
| Duffaud et al. 2021 | Low | Low | Low | Low | Low | Low |
| Le Cesne et al. 2023 | Some concerns | Low | Low | Low | Low | Some concerns |
| Duffaud et al. 2024 | Low | Low | Low | Low | Low | Low |

#### 3.4.1. Pooled analysis of PFS comparing regorafenib with the control groups

The evaluation of PFS was undertaken, employing the Response Evaluation Criteria in Solid Tumors (RECIST) across three distinct studies. Subsequently, a pooled analysis was executed to compare the PFS outcomes between the studies for both the regorafenib and the control groups. Cense *et al.’s* study was excluded because of insufficient data.

To combine the PFS findings across the aforementioned histological subtypes, a random-effects meta-analysis employing the inverse variance method was undertaken. Also, a subgroup analysis and test for subgroup differences were implemented. According to the results, regorafenib significantly improved PSF in bone sarcoma, with no evidence of heterogeneity across various types of bone sarcoma (MD = 9.69 weeks; 95% CI: 4.54, 14.84; I2 = 0%; P-heterogeneity = 0.87) **[Figure 1]**.

**Figure 1.**
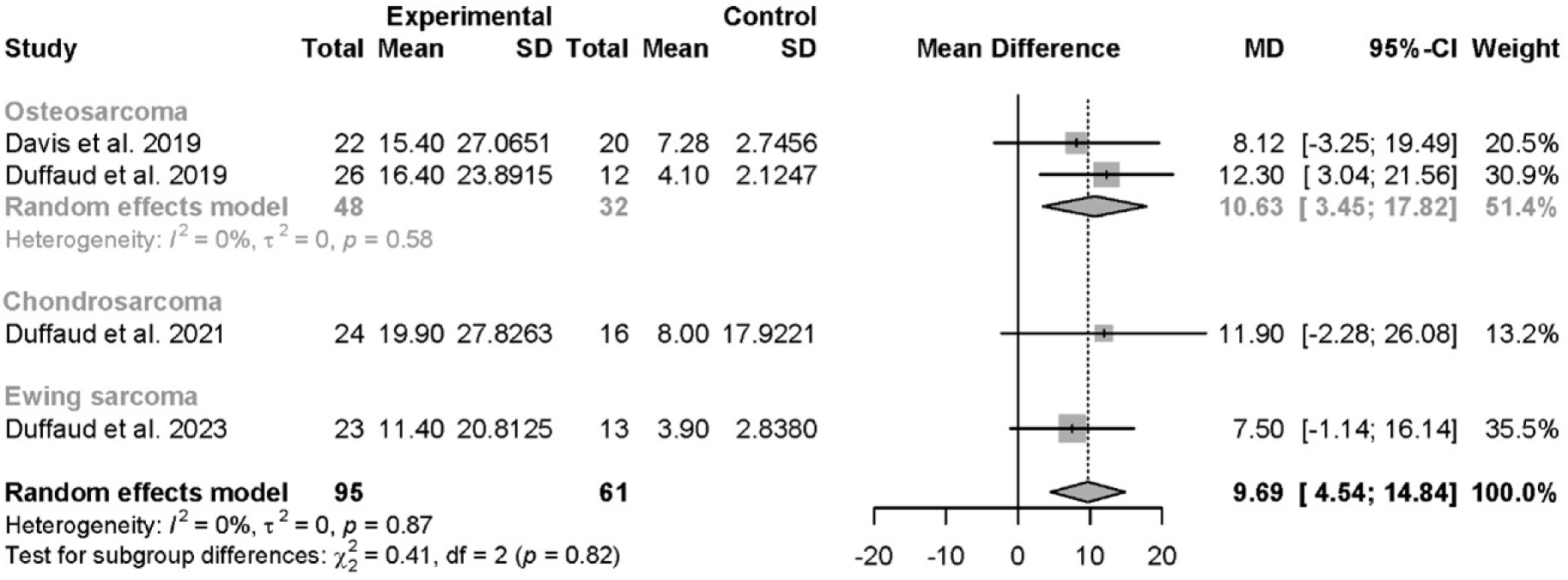
Meta-analysis of PFS comparing regorafenib to the control group.

#### 3.2.2. Pooled analysis of OS comparing regorafenib with the control group

OS was reported in all five RCTs. However, the Cense et al. study and Dauffaud et al. (2023) were excluded due to a lack of sufficient information. A pooled analysis was also conducted to compare OS between regorafenib and control arms across metastatic or locally advanced chondrosarcoma and metastatic osteosarcoma cohorts.

A subgroup analysis and test for subgroup differences were employed. According to the results, no statistically significant difference was observed between the treatment group and the placebo, with no heterogeneity. Additionally, the test for subgroup difference revealed no statistical difference between various types of bone sarcoma (MD = 0.85 weeks; 95% CI: - 36.33, 38.02; I2 = 0%; P-heterogeneity = 0.38) **[Figure 2]**.

**Figure 2.**
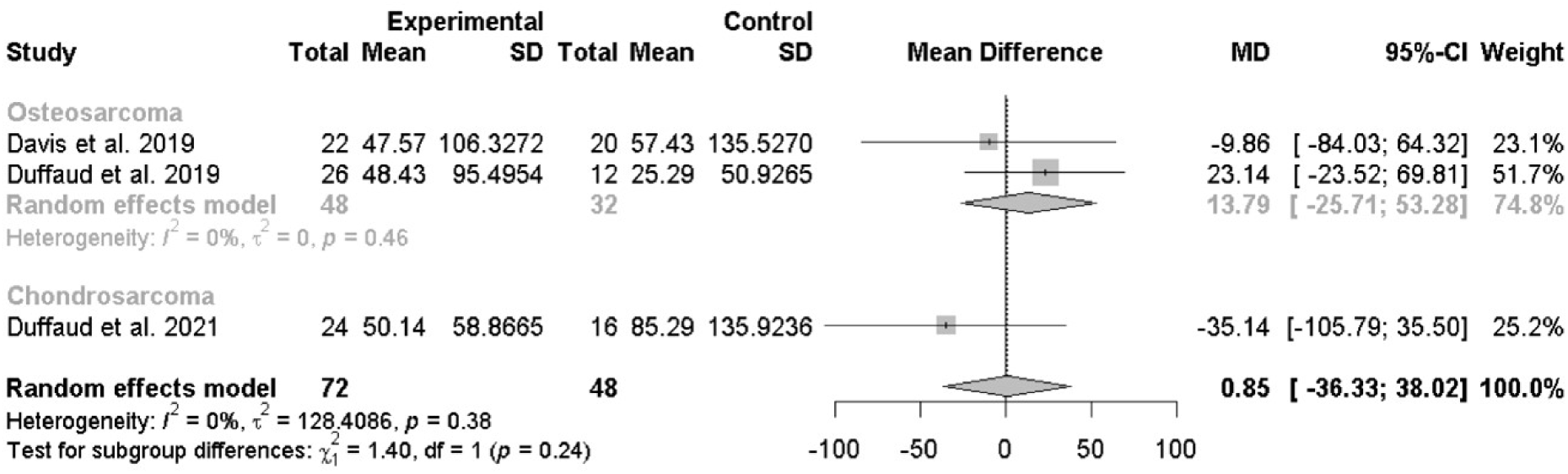
Pooled analysis of Progression-free survival comparing regorafenib with the control group.

#### 3.4.2 Pooled analysis of the safety profile comparing regorafenib with the control group

All publications were incorporated into the safety evaluation. All papers assessed safety using the National Cancer Institute’s Common Terminology Criteria for Adverse Events version four **[Table 4]**.

**Table 4.** Predominant Adverse Effects.

Regorafenib for Bone Sarcoma: A Meta-Analysis
| Study | Adverse effects |  |  |  |  |
| --- | --- | --- | --- | --- | --- |
| Davis *et al.* 2019 | Any grade | Hand-foot skin reaction (8/22) | Hypertension (7/22) | Nausea (5/22) | Diarrhea (4/22) |
|  | Grade three–five | Hypertension (3/22) | Maculopapular rash (2/22) | Thrombocytopenia (2/22) | Hypophosphatemia/Extremity pain (2/22) |
| Duffaud *et al.* 2019 | Any grade | Fatigue (26/29) | Hand–foot skin reaction (15/29) | Diarrhea (13/29) | Hypertension (12/29) |
|  | Grade three–five | Hypertension (7/29) | Fatigue (3/29) | Chest pain (3/29) | Hypophosphataemia/Hand-foot skin reaction (3/29) |
| Duffaud *et al.* 2021 | Any grade | Pain<sup>3</sup> (20/25) | Asthenia/fatigue (18/25) | Diarrhea (13/25) | Hand-foot skin reaction (12/25) |
|  | Grade three–five | Pain (5/25) | Hypertension (3/25) | Asthenia/fatigue (3/25) | Neuralgia/Thrombocytopenia/Diarrhoea (2/25) |
| Le Cesne *et al.* 2023 | Any grade | Asthenia (14/18) | Pain (12/18) | Hand-foot skin reaction (11/18) | Hypertension (10/18) |
|  | Grade three–five | Hypertension (4/18) | Hand-foot skin reaction (4/18) | Pain (4/18) | Diarrhea (3/18) |
| Duffaud *et al.* 2023 | Any grade | Pain (17/23) | Asthenia (16/23) | Diarrhea (14/23) | Hand-foot skin reaction (12/23) |
|  | Grade three–five | Pain (5/23) | Asthenia (4/23) | Thrombocytopenia (3/23) | Hand-foot skin reaction / Diarrhea (3/23) |

<sup>3</sup> The term "pain" was categorized under general disorders in the studies by Duffaud et al. (2023), Duffaud et al. (2021), and the CENSE study (2023). Additionally, these studies reported instances of skin pain and abdominal pain, which were not included in the overall counts for the term "pain."

Fatigue/Asthenia was the most frequent adverse effect, although it was not reported by Davis *et al.* Following this, the predominant adverse effects arising during treatment that were most documented consistently across all articles were hand-foot reaction(OR=9.30, 95% Cl=3.59-24.07, I^2^= 0%, P-heterogeneity = 0.69), diarrhea (OR=2.77, 95% Cl=1.28-6.01, I^2^= 0%, P-heterogeneity = 0.82), and hypertension (OR=4.00, 95% Cl=1.61-9.94, I^2^= 0%, P-heterogeneity = 0.39) **[Figure 3] [Figure 4] [Figure 5]**.

**Figure 3.**
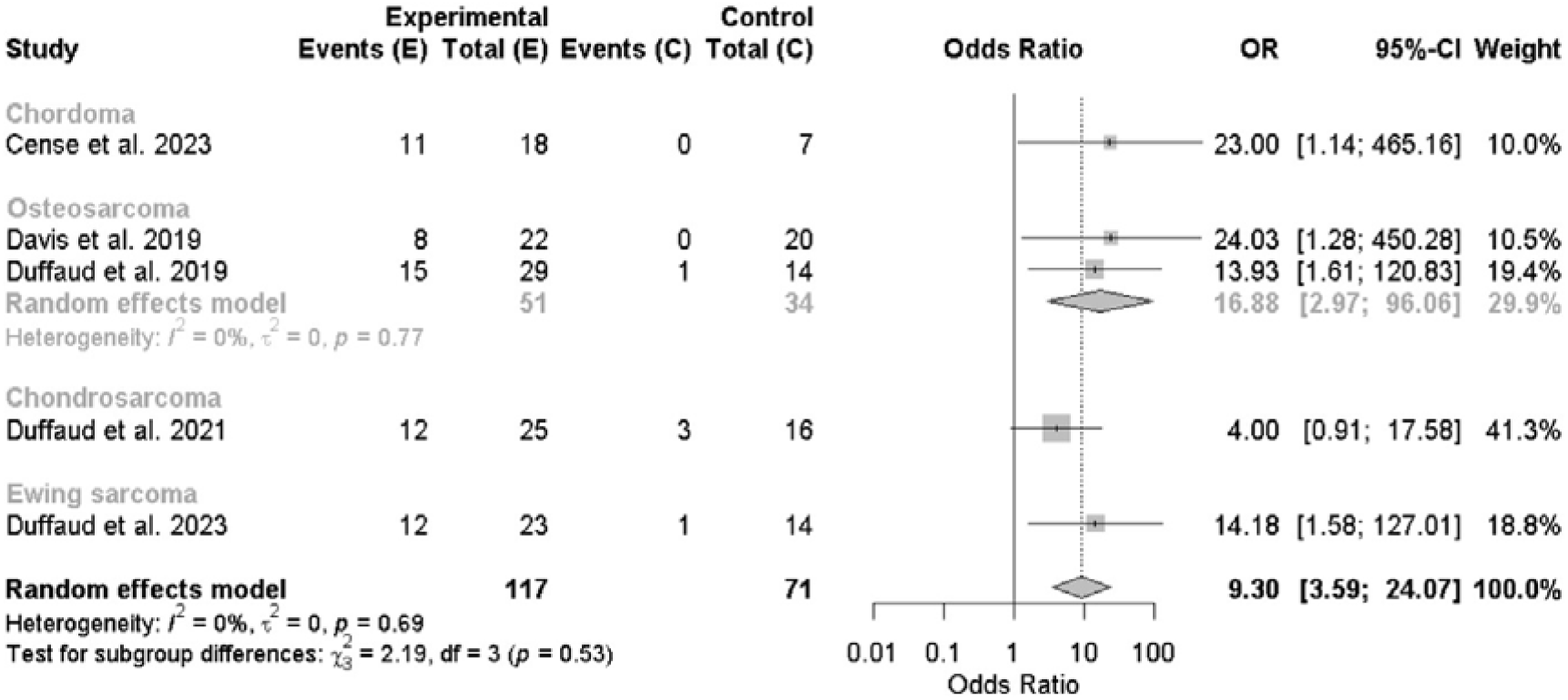
Pooled analysis of hand-foot reaction comparing regorafenib with the control group.

**Figure 4.**
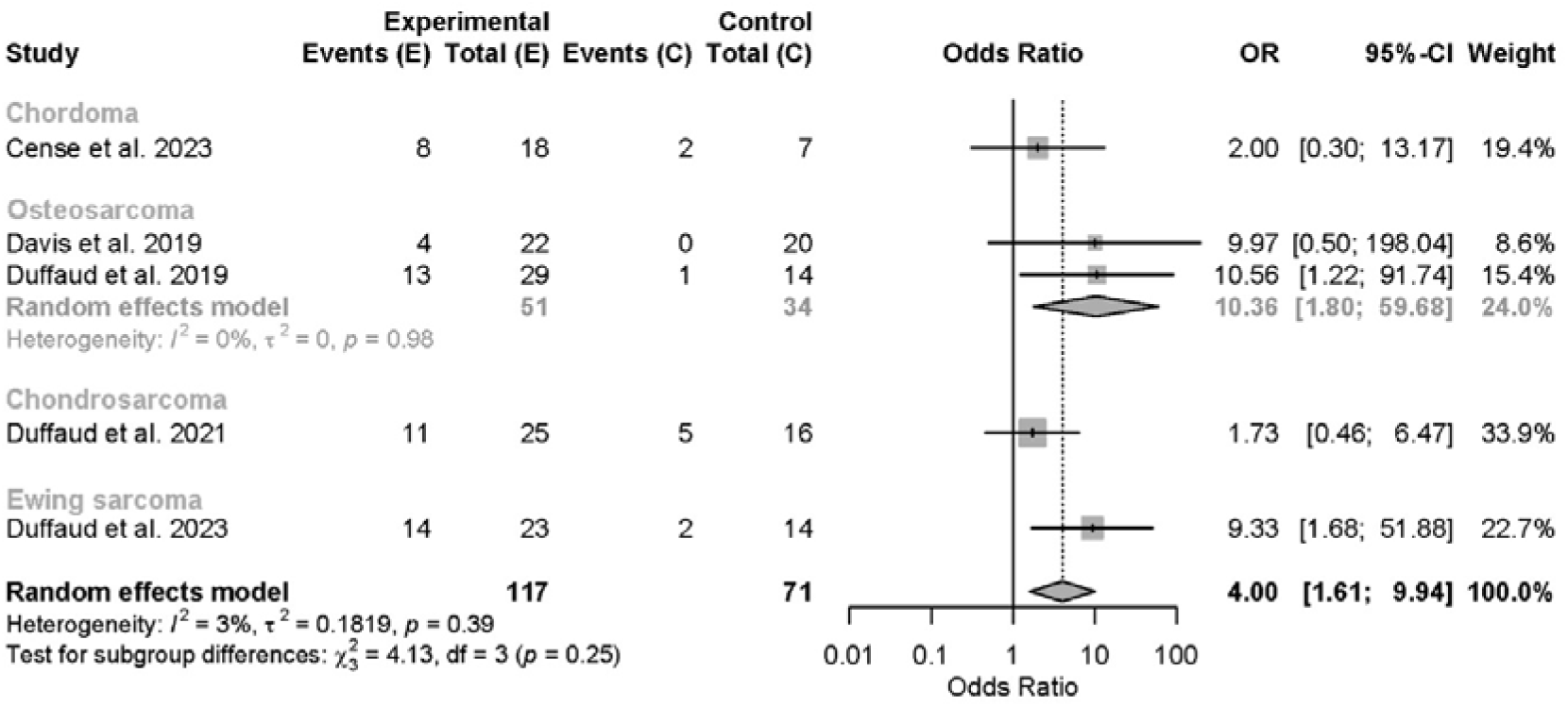
Pooled analysis of diarrhea comparing regorafenib with the control group.

**Figure 5.**
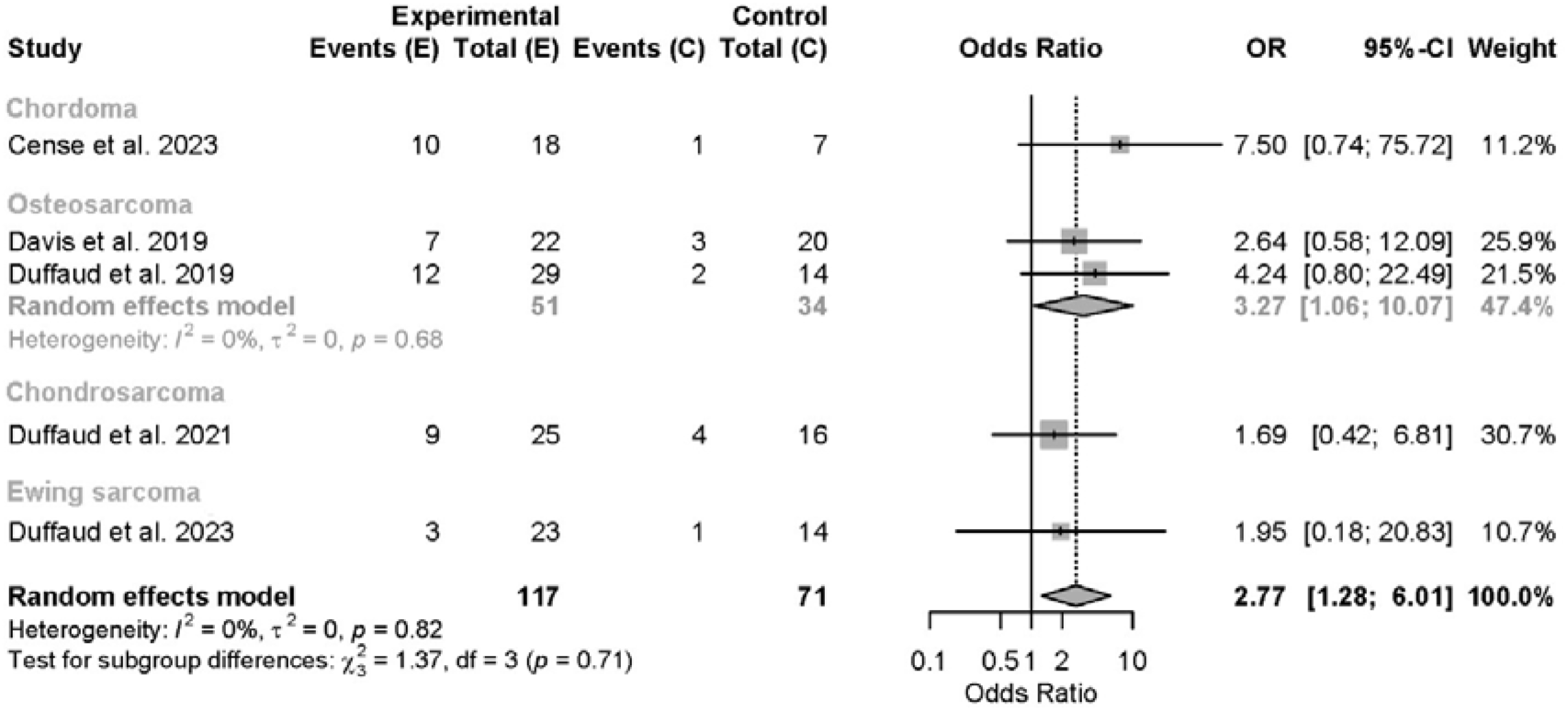
Pooled analysis of hypertension comparing regorafenib with the control group.

## 4. Discussion

### 4.1. Efficacy of regorafenib

Pre-clinical studies have shown that regorafenib exhibits broad anti-tumor activity, particularly against Ewing sarcoma, osteosarcoma, rhabdomyosarcoma, and doxorubicin-resistant Ewing sarcoma, as evidenced by its inhibition of tumor cell growth in both cell line and patient-derived xenograft models. (^23^) This systematic review analyzed five studies using progression-free survival as the primary endpoint, not objective response. Duffaud *et al.* argued that tumor shrinkage in calcified osteosarcoma lesions may misrepresent antitumor activity. Moreover, all studies used Response Evaluation Criteria in Solid Tumors version 1.1 (RECIST v1.1) for their outcome measurement. For efficacy analysis, the study conducted by Le Cesne *et al.* was excluded because insufficient information was available for analysis. (^24^)

The REGOBONE trial, a Phase 2, randomized study, showed a median PFS of 16.4 weeks with treatment versus 4.1 weeks with control group. Duffaud *et al.* used a modified intention-to-treat analysis but found a clinically meaningful PFS difference, with 65% non-progression at eight weeks vs. 0% in placebo. The David *et al.* study, part of SARC024, reported a median PFS of 3.6 months (treatment) versus 1.7 months (control), with 8- and 16-week PFS rates of 79.0% and 44.4% versus 25.0% and 10.0%, respectively. Both trials confirmed modest response rates, but regorafenib significantly improved PFS. (^18,25^) The OS was not statistically significant, possibly because of the crossover design.

Various tyrosine kinase inhibitors exhibit considerable effectiveness in treating osteosarcoma. A report encompassing 15 patients who received pazopanib treatment revealed a single instance of partial response (7%) and a median PFS duration of six months. (^26^) A phase II trial employing a single-arm design and utilizing sorafenib demonstrated a four-month PFS rate of 46%, with a median PFS of four months. Additionally, an 8% partial response (PR) rate was observed, involving three out of 35 cases, in a similarly designed trial involving apatinib, a four-month PFS rate of 57% was achieved, with a median PFS of 4.5 months. Notably, an impressive partial response rate of 43% was observed, involving 16 out of 37 patients. (^27^)

Preliminary results from a study investigating the use of lenvatinib in osteosarcoma were presented at ASCO 2018. These results indicated a four-month PFS rate of 33%, with a median PFS of 3.4 months. Additionally, an 8% PR rate was observed, involving two out of 26 cases. (^28^) In a recent open-label phase II trial investigating the use of cabozantinib, the results indicated a six-month PFS rate of 33%, with a median PFS of 6.2 months. Furthermore, a 12% PR rate was observed, involving five out of 42 cases. (^29^) While immune checkpoint inhibitors show limited efficacy in metastatic osteosarcoma, combining them with multi-kinase inhibitors like regorafenib, which targets VEGF receptors, has demonstrated synergistic effects, suggesting enhanced therapeutic potential across various cancers. (^30,31^)

### 4.2 Safety of regorafenib

Considering safety, treatment-related adverse events occurred frequently but were generally controllable and aligned with the known profile of multi-target tyrosine kinase inhibitors. (^35–38^) While hypertension was not the most common adverse effect, it emerged as the most commonly described grade ≥3 side effect of regorafenib in most of the studies included. Elevated blood pressure represents a recognized complication inherent to therapies targeting VEGF pathways, emerging as a direct pharmacodynamic consequence of intentionalmolecular inhibition. (^39,40^)

In Cense *et al.*, 68% of patients required dose reductions, and 18.7% discontinued treatment due to toxicity. Grade ≥3 adverse events included hand-foot reaction (22%), hypertension (22%), pain (22%), diarrhea (17%), with 18% experiencing serious treatment-related events.(^24^)

The high rate of dose reductions implies that regorafenib at the full dose of 160 mg/day may not be well tolerated. While toxicity led to discontinuation for three patients, 11 others required temporary treatment interruptions. A total of five serious adverse events attributed to treatment emerged among three individuals receiving treatment—comprising acute pancreatitis (n=1), cholecystitis (n=2), a pulmonary complication (n=1), and one instance of epilepsy documented in a placebo-treated participant, with no treatment-related death.

Davis *et al.’s* study demonstrated that regorafenib-related adverse events were more common with regorafenib. Dose reductions were required for 55% regorafenib patients, with the median dose at treatment end being 120 mg. Dose interruptions occurred for 13 (59%) on regorafenib. Beginning at a lower dose, such as 80-120 mg, may allow for longer treatment while maintaining efficacy. (^18^)

The specific adverse events, such as hand-foot reaction, hypertension, and diarrhea, are consistent with the known toxicity profile of regorafenib. However, the cases of severe pneumothorax (grade three) and colonic perforation (grade four) in responding patients highlight the potential for uncommon but dangerous complications. Due to AEs, these two individuals discontinued therapy after completing only two cycles at the 160 mg dosage, yet remained on treatment at reduced doses of either 120 mg or 80 mg. The study reported no Grade 5 AE or deaths.

Correspondingly, in the Duffaud et al. study, a starting dose of 160 mg/day regorafenib was administered. Serious treatment-related adverse events occurred in seven (24%) regorafenib patients, including severe hypertension, hypophosphatemia, hand-foot reaction, and others. (^25^)

No grade five events or deaths occurred, but frequent adverse events suggest the 160 mg/day dose may be excessive, warranting further dosing investigations. Personalized dose adjustments could enhance long-term tolerability. Side effect reporting varied across studies, particularly for fatigue and pain. In Duffaud *et al.* (2021), 18 out of 25 regorafenib patients reported fatigue, while 23 out of 29 did in Duffaud et al. (2019). Similarly, Cense *et al.* (2023) found that 14 out of 18 reported asthenia.

## 5. Conclusions

This systematic review and meta-analysis confirm the potential efficacy of regorafenib in bone sarcoma, significantly improving progression-free survival (PFS) with minimal heterogeneity. However, its impact on OS remains inconclusive, necessitating larger trials with extended follow-up. The analysis also highlights key limitations, including small sample sizes, restricted long-term safety assessment, and a lack of hazard ratios for PFS/OS, as most studies reported only point estimates. No pediatric patients were included, despite osteosarcoma’s prevalence in adolescents, emphasizing the need for pediatric-specific toxicity and dosing studies.

The safety profile indicates manageable but considerable toxicity, with hand-foot reaction, hypertension, and diarrhea as the most common adverse events. Frequent dose reductions suggest that 160 mg/day may be excessive for long-term use. Given the rarity of bone sarcomas, multi-institutional collaborations are essential to validate regorafenib’s role across sarcoma subtypes, optimize dosing strategies, and confirm long-term survival benefits.

## Data Availability

All data analyzed in this study were extracted from previously published clinical trial reports. No new datasets were generated. All relevant data are contained within the manuscript and its supplementary materials.

## Notes

### Competing Interest Statement

The authors have declared no competing interest.

### Funding Statement

This study was funded by a research grant from North Khorasan University of Medical Sciences (NKUMS). The grant was provided to support the research process and may be used to cover publication-related costs if applicable. No additional financial support or services were received from any third party.

### Author Declarations

The Ethics Committee of North Khorasan University of Medical Sciences waived ethical approval for this work.

